# Pilot Feasibility Clinical Trial of Virtual Reality for Pain Management During Repeated Pediatric Laser Procedures: Study Protocol for a Randomized Clinical Trial

**DOI:** 10.64898/2026.04.21.26351381

**Authors:** Megan Armstrong, Hannah Williams, Esteban Fernandez Faith, Ai Ni, Henry Xiang

## Abstract

**Background:** Lasers have wide applications in medicine and dermatology, but are associated with pain and anxiety, particularly in younger patients. Pain mitigation is often limited to topical anesthetics in the outpatient setting. Distraction techniques are limited by the need for ocular protection, which can include adhesive eye patches that can completely occlude vision. Virtual reality is effective at managing procedural pain and anxiety under other short medical procedures and is a promising tool for this population.

**Objective:** This trial aims to assess the safety, feasibility, and efficacy of Virtual Reality Pain Alleviation Therapeutic (VR-PAT) for pain management during outpatient laser procedures.

**Methods:** 40 patients requiring outpatient laser therapy for at least two sessions will be recruited from a pediatric hospital in the midwestern United States for this crossover randomized, two-arm clinical trial with a 1:1 allocation ratio. During the first laser visit, the participant will be randomly assigned to either play the VR-PAT game during their procedure or wear the headset with a dark screen. Participants will answer questions about their pain (Numeric Rating Scale (NRS) 0-10), anxiety (State Trait Anxiety Inventory for Children, NRS 0-10, Modified Yale Preoperative Anxiety Scale (mYPAS)), and pain medication usage. Those playing the VR-PAT will additionally report simulator sickness symptoms and their experience playing the game. At their second laser visit, participants will crossover to the opposite intervention from their first visit. The primary outcomes are the difference in self-reported pain and anxiety between the two interventions. Feasibility outcomes include the proportion of screened patients who are eligible, consent, and complete both visits and adverse events reported. To evaluate the efficacy of pain reduction, composite scores of pain score, pain medication will be calculated for each laser visit. To evaluate the efficacy of anxiety reduction, the change of mYPAS scores will be compared between control and VR groups at each visit using Wilcoxon rank sum tests. All statistical analyses will follow the intention-to-treat principle in regard to intervention assignment at each visit.

**Results:** The study was funded in January 2023 and began enrollment at that time. A total of n=44 participants were recruited and data collection was completed in November 2025, with n=40 subjects completing both visits. The sample was balanced with n=40 subjects using the intervention and participating in the control condition. The age range of the complete sample was 6 to 21 years at recruitment and was 55% female sex. Data analysis is in progress with final results planned for June 2026.

**Conclusions:** Findings from this innovative randomized clinical trial will provide early evidence on the efficacy of the VR-PAT for reducing self-reported pain and anxiety during outpatient laser procedures. The results from this trial will inform a large-scale, multisite study.

**Trial Registration:** ClinicalTrials.gov: NCT05645224 [https://clinicaltrials.gov/study/NCT05645224]

## Introduction

### Background and Significance

Lasers have wide applications in medicine and dermatology. In pediatric patients, laser technology is often used to treat congenital and acquired skin conditions. Pulsed dye laser (PDL) is the standard of care for treatment of vascular birthmarks, primarily port wine stains and capillary malformations. This laser is also used to treat infantile hemangiomas (residual changes and ulceration), spider angiomas, and pyogenic granulomas.

Additionally, lasers have become a critical component in the treatment of scars resulting from burns, trauma, surgery, and inflammatory conditions. Apart from vascular lasers, such as PDL, ablative lasers, such as the fractional CO_2_ laser, are used to improve the size, texture, symptoms, and function limitations caused by scars.

Though it is an effective and important treatment modality in pediatric dermatology, laser therapy is often associated with significant pain for the patient [1, 2]. Pain associated with laser procedures is typically described as heat or burn sensation. PDL is described as a hot rubber band snapping on the skin with each pulse [3] and the laser pulses 15-100 times per treatment session [4]. Infants and young children may require some form of anesthesia [3], and even adolescents and adults may have difficulty tolerating the procedure [5].

It is vital to mitigate pain and anxiety during laser procedures in pediatric patients. Unrelieved pain during pediatric procedures has been associated with adverse effects on recovery and postoperative behavioral abnormalities [6–8] including post-procedural symptoms of post-traumatic stress disorder [9]. Anxiety and pain can lead to a lack of cooperation with the procedure by the patient, which may impact the effectiveness of the therapy [5]. Darkness from laser eye protection and pain-related distress may also precipitate phobic responses in the patient [4], which increases their anxiety and can negatively impact subsequent treatments [10] as well as future interactions in healthcare settings [11]. Experiences of undertreated pain can have significant adverse effects on immediate and long-term mental health for pediatric patients [12]. Therefore, effective pain mitigation during pediatric laser procedures is a crucial component of biobehavioral intervention targeting pediatric patients.

Pharmacologic analgesia is often the primary pain management technique during pediatric laser procedures. Topical anesthetics, often lidocaine, are the most common analgesics chosen during pediatric laser procedures [13]. Topical agents are generally well-tolerated in the pediatric population and can reduce pain associated with laser procedures, but they are often insufficient for pain mitigation in all patients [3]. Also, topical agents may not be a viable option when the lesion involves large regions of the body or is located near the ears, fingers, eyes, and nose [2]. Local injectable lidocaine is an additional pain relief option. However, epinephrine is typically included in the injected solution, which may decrease lesion size and hinder the effectiveness of the laser treatment for vascular lesions [3]. Furthermore, use of a needle may result in increased pain or discomfort, lesion distortion, and there may be concern for lidocaine toxicity if the anesthetized area is large [5]. General anesthesia is an option that is occasionally utilized [13], and it can be especially useful for highly uncooperative or distressed children [3]. However, this option is very carefully considered due to growing concern for neurocognitive defects resulting from the use of general anesthesia in children [13, 14]. Sedatives, often midazolam, can be administered to alleviate both pain and anxiety, as well as to induce anterograde amnesia [5]. Unfortunately, midazolam is associated with a primary side effect of respiratory depression, which is of serious concern [3]. It is preferable to treat dermatologic conditions as early as possible [5]; the response rate to laser procedures is significantly better in younger children [15] and delaying the procedure may negatively impact psychosocial development due to exposure to social stigma [4]. Therefore, waiting until the child has aged to maximize tolerance of the laser procedure is not ideal. Efforts must be made to minimize the pain and anxiety of the laser procedure for the pediatric patient.

Ocular exposure to laser radiation can result in permanent eye damage, including vision loss [5]. It is paramount that any eye protection used during the procedure can effectively block radiation exposure to the patient’s eyes. A variety of eye protections can be used. The eye patches fully cover the eye and thus submerge the patient in darkness, which can induce anxiety during these laser procedures. This special eye protection needed by patients during laser treatment creates a unique opportunity to use a virtual reality (VR) headset not only as eye protection but also as a pain and anxiety management strategy.

### Gap in Knowledge

There is a clear need for the development of alternative nonpharmacologic pain and anxiety relief modalities for pediatric procedural pain. Recently, distraction has emerged as a highly effective pain-mitigation tool for this population. According to the Cognitive-Affective Model of Pain, distraction alleviates pain by consuming attentional resources that would otherwise be devoted to pain processing [16], thereby decreasing subjective pain and related distress [17]. Various forms of distraction have been found to be effective at reducing subjective pain [10, 18], decreasing pain-related brain activity [19] and increasing positive procedural outcomes [20] in the pediatric population.

Among the various forms of distraction, VR has been found highly effective in medical settings for pediatric patients. VR utilizes a head-mounted display to create a highly immersive environment that blocks the sights and sounds of the hospital and can distract the user from real-world pain [11]. VR has been found to be significantly better than the standard of care at decreasing self-reported pain [21–23], anxiety [10, 24], time spent thinking about pain [10, 23], heart rate [18] and pain-related brain activity [25, 26] during painful pediatric procedures. VR has also been found to improve cognitive and emotional processing of the event [18] and VR use reduces the formation of phobic responses to hospital environments [27].

The vast majority of these studies reported no instances of simulator sickness or other immediate adverse effects of using VR [10, 25, 28] and reported that the majority of the patients found the VR experience to be enjoyable [23, 28]. In addition, both parents and nurses tend to be satisfied by the experience of using the VR on the patient [29] and nurses have reported the children are more cooperative during the procedure while using VR, which improves the efficacy of the procedure itself [12]. Additionally, the effectiveness of VR-based pain relief does not seem to diminish when used repeatedly, and patients continue to find the technology fun to use [23, 27]. VR is a promising anxiety and pain relief modality that is becoming increasingly flexible, immersive and affordable, making it easy to use in medical settings and highly applicable to a wide variety of patients and medical settings [28].

Despite extensive research regarding VR use in medical settings, there remains a knowledge gap regarding the efficacy and safety of VR in pediatric laser procedures. Most previous studies were conducted in the setting of burn wound dressing changes, IV placement or lab-induced pain. We are only aware of very few studies of the use of VR in the setting of laser procedures. Jaquez et al. found promising early results for non-pharmacologic distraction during a variety of dermatologic procedures, but this was not exclusive to PDL or CO_2_ lasers [30]. More research is needed to understand the ability of VR to be applied in this setting. Additionally, little is known about the safety of using VR headsets during laser procedures. Ocular exposure to laser radiation can result in permanent eye damage, including vision loss [5]. It is paramount that any eye protection used during the procedure can effectively block radiation exposure to the patient’s eyes.

### Prior Work

Our previous clinical study provided strong evidence about the efficacy of the smartphone version of the virtual reality pain alleviation therapeutics (VR-PAT) for significant pain reduction during burn dressing changes [29]. This randomized clinical trial (RCT) tested the efficacy and feasibility of this tool among 90 pediatric outpatient burn patients aged 6 to 17 years. Participants were randomly assigned to an active VR-PAT (N=31), passive VR-PAT (N=30), or a standard care (N=29) group during a burn dressing change. Active VR-PAT significantly reduced observed and self-reported pain during burn dressing changes. Patients/caregivers reported satisfaction with the VR-PAT, and nurses reported the tool could be easily used in clinics.

In another RCT study, we examined the efficacy of VR-PAT as a pain alleviation tool during at-home dressing changes for pediatric burn patients (5-17 years) [31]. Participants (n=35) were randomly assigned to either the active VR-PAT or the control condition. Subjects found the VR-PAT to be a useful distraction during home dressing changes and reported it to be easy to implement. In the VR-PAT group, children and caregivers reported that pain decreased as the week of dressing changes progressed and was lower than that in the control group after the fourth dressing change. Children playing the VR-PAT reported consistent happiness and fun as the week went on alongside increased realism and engagement, indicating that repeated use of the tool does not diminish its efficacy.

Our previous studies and preliminary results position us well to expand the VR-PAT for anxiety relief and pain reduction during pediatric laser therapy.

### Objectives

The overall objective of this proposal is to conduct a randomized crossover clinical trial to assess the safety, feasibility, and efficacy of VR-PAT for pain management during outpatient laser procedures and to assess anxiety as a preprocedural key factor for pain during procedures. Our specific aims are to 1) assess the feasibility and efficacy of VR use to reduce pain in patients undergoing dermatologic laser procedures and 2) evaluate the impact of VR use on reducing anxiety in patients undergoing dermatologic laser procedures. Our central hypothesis is that VR-PAT is safe to use and can effectively and significantly reduce pain during pediatric dermatologic laser procedures.

## Methods

### Trial Design

We will conduct a prospective, two-group crossover RCT with a 1:1 allocation ratio to either the intervention (VR-PAT gameplay) or the control (VR headset turned off) for the first laser appointment. Participants will crossover to the opposite group at their second laser appointment. Pain, anxiety, and VR engagement (for those in the intervention group) will be assessed after each research appointment, spaced about six to eight weeks apart. The flow of participants can be seen in **Figure 1**. The findings will be reported based on the CONSORT (Consolidated Standards of Reporting Trials) guidelines.

### Settings and Participants

Participants will be enrolled from the outpatient dermatology clinic on the main campus at Nationwide Children’s Hospital (NCH) in Columbus, Ohio. We plan to recruit n=40 patients to finish two laser visits (age 5+ years old) who clinically require laser therapy. The inclusion criteria are 1) dermatology patients (5+ years) who are undergoing the first of a new series of laser procedures at the NCH Outpatient Dermatology clinic; 2) have a legal guardian present for patients less than 18 years old for the procedure (for informed consent); and 3) can communicate orally. Exclusion criteria include 1) any wounds that may interfere with study procedures; 2) usage of the diode laser; 3) vision, hearing, or cognitive/motor impairments preventing valid administration of study measures; 4) history of motion sickness, seizure disorder, dizziness, or migraine headaches precipitated by visual auras; 5) minors in foster care; 6) unable to communicate in English; 7) pregnant women; or 8) prisoners. Exclusion of diode laser use was determined during safety testing, in which laser safety was confirmed for PDL and CO_2_ lasers, but not diode lasers. Laser safety testing was performed by a third-party company to test the VR headset for ocular safety. They determined that this headset was safe to use for PDL and CO_2_ lasers but did not protect against diode laser wavelengths. Based on our previous studies, patients under five years old were unable to respond to pain measurement and other study questionnaires effectively.

The laser team (led by our physician champion) will identify potential study subjects, and our research associates will screen them for eligibility. Once a patient’s eligibility is confirmed, a trained researcher will approach the patient and their legal guardian (if necessary) to introduce the study. Trained research staff will ensure subjects are awake, alert, and able to provide assent (if child >9 years but <18 years old) or consent if age 18+ and demonstrate understanding of the study before being consented. According to the NCH IRB policy, patients less than 9 years old only need consent from a caregiver who is the legal guardian when the study involves only minimal risk, however all subjects will be asked for verbal assent. Signed consent and assent (if applicable) will be obtained before formally enrolling patients into the study. During the informed consent conversation, the legal guardian and the patient (if age 18+) will be asked for permission to store the subject’s protected health information (PHI) and identifiable information for future IRB-approved research. The person providing consent can opt out of this option, which will not affect their standard of care treatment. Biological specimens will not be collected for this study.

### Participant Timeline

Study enrollment at NCH – patients will be recruited prior to their first sequential dermatologic laser procedure. After consent and assent (if applicable) are obtained, patients will be randomized to either the VR-PAT intervention or control. A research associate will collect background information, such as demographics, anxiety, and prior video game experience, prior to randomization. VR-PAT instructions will be provided to the intervention group prior to participation.

Laser procedures at NCH (n=2 visits) – patients will receive their laser procedure as usual. Participants in the intervention group will wear the Pico headset and play the VR-PAT during their procedure. Participants in the control group will receive their laser procedure while wearing the VR headset (without the game). A researcher in the laser treatment room will observe the procedure and will document for the intervention group, the amount of time playing the VR-PAT, whether the participant declined to use, and number of interruptions of VR. Following the procedure, a different researcher (blinded to the group) will ask both groups of participants about their pain experience and satisfaction with the game. This will be repeated once for each participant as they will cross-over to the opposite intervention group at their next laser procedure.

### Randomization and Blinding

A block randomization scheme with a block size of four will allocate participants to one of the two intervention sequences (VR-PAT followed by control or control followed by VR-PAT) at a 1:1 ratio without stratification. We chose a block randomization strategy to avoid imbalances in allocation to the intervention at the first visit in the case we were unable to recruit our planned sample. The randomization scheme was developed and uploaded to a study-specific Research Electronic Data Capture (REDCap) project by the project manager. Study participants are randomized into intervention sequences after signing consent and completing the baseline survey. Research recruiters and subjects do not know the intervention assignment until the subject is randomized in REDCap.

We cannot blind the participants or the clinical team to the assigned intervention since the headset will either be turned on or off. The researcher who randomized the participant and observed the laser procedure will not be blinded, but a second researcher who is blinded to the intervention group will collect pain ratings and anxiety post-procedure. The second researcher will wait in a different room during randomization and the laser procedure, and the first researcher will remind the participant at the end of the laser procedure to keep their group a secret until they are asked. The study biostatistician will also be blinded to the intervention group while performing analyses on the primary outcome. The data will be blinded by removing labels such as “VR” and “Control,” and replacing them with unidentifiable letters (e.g., “A” and “B”). The biostatistician will need to be unblinded for secondary outcomes, as only participants in the intervention group will answer the VR experience questions.

The blinded researcher will need to be unblinded after pain and anxiety ratings are obtained, as those in the control environment will not be able to answer the VR experience questions. The researcher will become unblinded by asking participants whether they played the game. The biostatistician will be unblinded for full analyses by providing the full dataset with complete data labels and VR questions. This unblinding will occur after the primary outcome analysis has been completed.

### Intervention

VR-PAT is the intervention for this RCT. VR-PAT is a standalone mHealth tool developed at NCH and does not require an internet or WiFi connection. The intervention is hosted on a Pico Neo 3 Pro Eye, which is a standalone VR headset that supports native eye tracking, enabling applications to gather real-time user feedback for behavior-based application control. Our VR game is the only application available on the headset and starts immediately upon pressing the power button, which will ensure all participants play the VR intended for this study. During the laser procedure, subjects will play the virtual game by slightly tilting their head, thereby minimizing interference with the procedure while cruising on a boat and aiming for the snow-blowing statues floating along the riverbank. The statues will burst and emit snow if the subject correctly aims at them, and a thermometer placed in the front of the boat will show decreased temperatures as more snowflakes are blown. As feedback to reinforce continued engagement, a scoreboard placed beside the thermometer will show subjects the number of statues he/she has activated. Additionally, as the temperature drops, snow and ice will start piling up on the boat and its surroundings, providing an enhanced “cooling” experience for dermatology patients. Audio from the headset provides surrounding and directionally-adaptive effects to match the progress of the game and the direction to which a subject’s head is turning, further enhancing the immersive experience of VR-PAT during dermatologic laser procedures. The active VR-PAT game can last indefinitely, so it can be used for the entire procedure without being interrupted.

Participants playing the VR will be advised to discontinue using the headset during a procedure if it causes motion sickness, headaches, nausea, or dizziness. The dermatologist will pause the laser procedure and switch the eye protection to one available in the outpatient clinic. Any adverse events from participation will be noted and reported on the subject surveys. The comparator or control for this study is the same VR headset without the game, just the dark screen. This comparator was chosen to maintain the experience of wearing something on the face for both groups. Patients undergoing a laser procedure are required to wear eye protection, which can include adhesive eye patches that occlude the eyes and would create a similar dark environment.

Participants under either intervention will be allowed to use any pain or anxiety medication the dermatologist deems appropriate. Any medications provided will be documented.

### Measures

All data will be entered directly into the REDCap database as it is collected. A second researcher will check the REDCap records after each visit, ensuring the completeness and accuracy of the data. Data collected for this study include:

<u>Participant Data</u> (self-reported): Prior to randomization, participants will report their prior video game experience (type of video games played, frequency, and favorite games). Before the laser procedure, all participants (control and intervention visits) will self-report their anxiety state using the State Trait Anxiety Inventory for Children [32] (STAI-CH; ratings from not at all to very much) and their current anxiety (Numeric Rating Scale (NRS), 0-10) [33]. After the procedure, the participant will answer the remaining survey questions assessing overall pain score (NRS, 0-10) [34, 35], worst pain score (NRS, 0-10), time spent thinking about pain (NRS, 0-10), length of the procedure (in minutes), and anxiety during the laser procedure (NRS, 0-10). Those playing the VR-PAT game will report their experience with the VR game (degree of realism, fun, engagement, and satisfaction; NRS 0=10) and qualitative questions about preferences with the VR-PAT. Participants are also asked to report any simulator sickness associated with the VR game and whether they would want to use the VR again for future laser visits outside of research. These data will be collected at each of the two laser procedures study visits.

<u>Legal guardian data</u> (self-reported when participants are <18 years): Before the laser procedure, all legal guardians will self-report their anxiety state using the short form State Trait Anxiety Inventory (STAI-6) [36]; ratings from not at all to very much). After the procedure, the legal guardian will report their perception of the participant’s pain by overall pain score (NRS, 0-10) and worst pain score (NRS, 0-10). These data will be collected at each of the two study visits.

<u>Nurse data</u> (self-reported): to assess the practicality and utility of the VR experience as a pain and anxiety management tool for dermatologic laser procedures, the nurses will report whether the distraction tool is helpful and if it is easy to use during laser procedures (Yes/No).

<u>Observed measures</u> (research collected): the Modified Yale Preoperative Anxiety Scale (mYPAS) [37] will be assessed at each laser procedure before entering the treatment room, before the procedure begins, and immediately following the procedure. This is a 5-item scale with a score of >30 indicating anxiety. During the procedure, the researcher will document the start and stop times of the laser procedure, the start and stop times of the VR (if applicable), whether the participant declined to use the VR-PAT, and the number of voluntary interruptions of the VR-PAT.

<u>Medical record review</u> (research abstracted): Researcher will review the participants medical record and document and pain-related medication (name and dosage) the participant used prior to the laser procedure. Demographics and procedure information recorded will include: date of birth, sex, race, ethnicity, and dermatologic variables (laser type, diagnosis, lesion size, lesion type, and lesion location).

#### Feasibility Outcomes

The feasibility of using VR during outpatient laser procedures will be evaluated by the proportion (%) of patients screened who are eligible to be approached, those who consent to the study, and those who complete both study visits. This will be tracked in our screening log and analyzed periodically throughout the study for funding reporting. Adverse events, including simulator sickness, will also be used to evaluate feasibility (see Adverse Events and Oversight section for specifics). The threshold for scaling up this study is no serious adverse events reported, less than 10% loss to follow-up due to the VR, and greater than 80% of both patients and nurses are satisfied with the intervention. We will not count clinical loss to follow-up in this measure. There is no pre-defined proportion for recruitment and consent, but these outcomes will be used to determine the possible sample size.

#### Primary Outcomes

Difference in self-reported worst pain, average pain, and time spent thinking about pain (NRS 0(min)-10(max)) between VR-PAT and control visits. These questions are asked at each visit and will be compared between the intervention and control visits.

Change in procedural anxiety (Modified Yale Preoperative Anxiety Scale (mYPAS) (observed by researcher), 23.33(min)-100(max), with higher scores denoting higher levels of anxiety).

Assessed at three time points (prior to entering procedure room, prior to the procedure, and immediately following the procedure) for each laser procedure. These differences will be compared before and after the procedure, and between the intervention and control visits.

#### Secondary Outcomes

Average self-reported VR experience (NRS 0(min)-10(max) for the degree of realism, pleasure, and satisfaction with VR). These VR experience questions are assessed immediately following the laser procedure at the intervention visit.

Nurse-reported utility (Binary (Yes/No) questions of whether the nurse found VR-PAT to be helpful and easy to use during the procedure) is assessed immediately following the laser procedure at the intervention visit.

### Adverse Events and Oversight

Participants playing the VR-PAT game will be asked to answer the question, “Did the game make you feel not well? If yes, please explain.” Study coordinators will carefully review collected data for any adverse events reported during the study intervention and immediately report this information to the project manager. The PIs will determine how serious the event was, whether it was related, and if it was unexpected. All serious adverse events will be reported immediately to the sponsor and IRB. At recruitment, participants will be informed of what simulator sickness symptoms to be aware of and advised that if they occur, we can pause the laser procedure to remove the headset and use the standard eye protection available. All participants will also receive a direct phone number for the research PI to report adverse events.

This trial will be overseen and coordinated by the co-PIs and a project manager. We will have at least one researcher to assist with the screening, recruitment, consent, and data collection. The research PI and project manager meet weekly to discuss the project, including recruitment updates, challenges, and any reported adverse events. There is no specific steering committee for this study.

### Sample Size

Since this is a pilot study, the focus of the statistical analysis will be estimating the effect sizes of VR-PAT and their 95% confidence intervals, as well as the variances of the outcome measures. These findings will facilitate sample size calculations and study power estimates for future larger-scale randomized trials. We decided on a sample of 40 participants (over two visits) based on the grant budget and the patient volume for the one dermatologist collaborating on this study. Across acute pain studies, clinically meaningful improvements are generally larger than a 1-point change on the 0–10 NRS, with systematic review evidence suggesting minimum clinically important differences of approximately 13–36% reduction (median ∼23%), corresponding to roughly 1.4–2.3 points on the NRS [38, 39].

### Data Analysis

To achieve Aim 1, composite scores will be calculated for each laser procedure that encompass pain score, pain medication use during procedure, and whether rescue medication was used. The rationale for choosing the composite score as the outcome is that prior researchers have shown a significantly high positive relationship between pain medication use and self-reported pain score [40]. From clinical perspectives, pain scores alone are influenced by patient actual opioid use and may not fully reflect treatment benefit of VR [41]. The composite score approach therefore provides a single, patient-centered outcome that represents overall analgesic benefit, aligning with the goal of achieving adequate pain control while minimizing opioid exposure. In our analysis, we will apply the rank-based approach by Dai et al. to assign weights to pain and medication use, reflecting their complementary clinical importance as measures of pain reduction [41]. In the primary analysis, the composite scores from the laser procedure with VR will be compared to those from the procedure with standard care (i.e., control) by paired t-tests or Wilcoxon signed rank tests (depending on the distribution of the composite scores) due to the cross-over design. The clinical effect size will be the mean difference (if paired t-tests are used) or median difference (if Wilcoxon signed rank tests are used) in the composite score between VR and control procedures. Standard deviation of the difference will also be calculated.

For Aim 2, the mYPAS score had 6 repeated measurements over time (T0, T1, and T2 during each of the two laser procedures). At the first procedure, we will compare the change of mYPAS scores from T0 to T1 and T0 to T2 between control and VR groups using either two-sample t-tests or Wilcoxon rank sum tests (depending on the distribution of the change of mYPAS scores). The clinical effect size will be the difference in medians of change of mYPAS, with the confidence limits calculated by bootstrapping with replacement with 1,000 repetitions.

Bootstrapping has been found to provide better confidence limits with asymmetric data, such as the mYPAS score in our study and is the recommended methods. The same statistical analysis approach has been successfully used by other researchers who reported that VR effectively reduced the preoperative anxiety among 71 children 5-12 years of age scheduled for elective surgery [42]. At the second procedure, we will compare the mYPAS scores at T0 and T1 between control and VR groups using Wilcoxon rank sum tests since mYPAS scores are shown to be asymmetric [42]. The clinical effect size will be the difference in medians of mYPAS. Confidence limits will be calculated using bootstrapping again.

Linear mixed-effect models with subject-specific random intercept will be fit on the composite scores to evaluate any modification of effects of VR on pain by sex, age, and anxiety with adjustment for baseline covariates and potential carryover effects of VR from the first to the second procedure. The following covariates will be included in the regression models: study group (VR vs control), period (2nd procedure vs 1st procedure), study group by period interaction (test for carryover effect), age, age by study group interaction, sex, sex by study group interaction, procedural anxiety, procedural anxiety by study group interaction, race, and ethnicity, STAI-CH, STAI-6, and average mYPAS scores between T0 and T1. We will include each of the three interaction terms in a separate regression model. Due to the limited sample size, testing the interaction may not be feasible. We will then run the main linear models without interaction terms. Our results will give us preliminary data for us to estimate the future study sample size and power. In case a significant carryover effect is detected, we will only use data from the first procedure to evaluate the VR effect using two-sample t-test or Wilcoxon rank sum tests (depending on the distribution of the composite scores).

All statistical analyses will follow the intention-to-treat principle, where participants will be analyzed according to their assigned study groups, regardless of compliance status and group switching during the study.

### Ethical Considerations

The NCH IRB approved this study (STUDY00002880) initially on October 17, 2022, and protocol version 6 was approved on April 1, 2025. Important protocol modifications have been communicated to study personnel by the project manager and will continue to be communicated. Written, informed consent to participate will be obtained from one parent or legal guardian of all participants <18 years of age and from the participant if age 18+, and written assent will be obtained from participants aged 9 years and older, as per the NCH IRB policy for written assent. This trial has been registered on ClinicalTrials.gov (NCT05645224), where detailed information about the study protocol, inclusion and exclusion criteria, interventions, outcomes, and ethical approval can be found. The trial registration process was completed before the enrollment of the first participant.

## Results

This study was funded by an intramural grant from the Clinical and Translational Science Institute at Nationwide Children’s Hospital (GRANT #: IFPAWRI012023) in January 2023. The enrollment started in January 2023 and data collection was completed in November 2025. A total of n=44 patients were recruited and completed the first visit, with n=40 subjects completing both visits. The sample was balanced with n=40 patients using VR intervention and n=40 patients participating in the control group. Reasons for not completing both visits included not continuing laser treatment (n=3) and not being interested in playing the VR game (n=1). Of those who completed both visits, the age range of the sample was 6 to 21 years at recruitment and was 55% female sex. The data analysis is expected to be finished in June 2026, with a manuscript to follow. Future results will be disseminated through publications in peer-reviewed journals and abstracts submitted to conferences in relevant disciplines.

## Discussion

Our VR-PAT protocols have been successfully implemented in our prior and ongoing studies, and we believe this protocol will be successful in a new setting. Prior VR research has shown to significantly reduce self-reported pain [21–23], anxiety [10, 24], time spent thinking about pain [10, 23] during painful pediatric procedures. We anticipate similar findings in this outpatient laser study. We also anticipate very few reports of simulator sickness since this study uses the same VR-PAT we have used previously during pediatric burn dressing changes [29, 31], the emergency department [43], and pin-pulling procedures [43]. We anticipate this intervention to be feasible to implement, with nurses reporting a high degree of helpfulness and ease of use, like in our previous work [29].

The strength of this study is that laser procedures are unique clinically, in that the pain stimulus is the same for each procedure. This allows us to use a crossover study design and within subject comparisons. Our prior VR research has been conducted during burn dressing changes, which are subject to healing over time, making each dressing change a different pain experience. Another strength of this study is providing pain and anxiety distraction during an outpatient procedure where that has been difficult. To maintain laser safety, shiny surfaces must be limited to avoid a reflection of laser beam and ocular safety requires glasses that filter the light waves and can alter the colors patients can see. Since our VR headset passed laser safety testing, it can be a valuable tool for this patient population. Finally, we have found that the proportion of participants recruited is high among those we have approached and retention has been high among those patients who returned to the clinic for their second laser visit. Part of our retention success can be attributed to the study’s buy-in from the clinic nurses. They have helped to keep excitement high among participants by talking to them about the VR game during the laser procedures, and they have even developed an anonymized scoreboard for participants to keep track of their scores. The nurses also keep the VR headset charged and clean to keep the study implementation running smoothly.

This study is subject to some known limitations. First, all the subjects come from the same hospital and all the laser procedures are performed by the same physician, which does limit our ability to generalize the findings. Second, since complete blinding was not possible due to the nature of the intervention, there is the chance of an expectation bias by participants and clinicians. The research team attempted to manage subject expectations at recruitment and prior to each visit by explaining the research nature of the VR. We also made the choice not to measure subject expectations for the VR because we felt that this would prompt a reflection of their expectations and peak excitement. The third limitation is in evaluating the possible sample site. There has been more loss to follow-up than anticipated due to patients who are not following up clinically. This limits our ability to retain them in this research study. The proportion of eligible subjects whom we recruited is also lower than expected. This is largely related to many screened patients not ultimately scheduling a laser procedure. This will be a consideration for future studies and determining the number of physicians and/or sites needed to complete a powered study.

Our future research direction following the successful completion of this study depends on our results but could include a fully powered efficacy study and eventually a multisite implementation study. In designing future implementation and dissemination work, sustainability will be a priority. This will include evaluating insurance reimbursement to cover the cost of owning and utilizing this VR. We would follow in the footsteps of companies like AppliedVR, which was the first VR to obtain FDA-authorization established a unique Healthcare Common Procedure Coding System (HCPCS) Level II code [44].

In conclusion, patients needing outpatient laser therapy have unique needs for pain and anxiety management, particularly in the context of ocular protection from laser damage. Virtual reality is a promising digital technology that can improve the experience of patients requiring dermatologic laser therapy. We anticipate that findings from this innovative randomized clinical trial will provide early evidence on the efficacy of the VR-PAT for reducing self-reported pain and anxiety during outpatient laser procedures.

## Supporting information

SPIRIT Checklist

## Acknowledgements

We would like to thank the nursing staff in the Division of Dermatology for their support of this project.

## Data Availability

The datasets generated and/or analyzed during the current study are not publicly available due to them containing PHI, but are available on reasonable request. To request access to the de-identified research data, please contact the Data Trust & Value Committee at Nationwide Children’s Hospital via e-mail at. Deidentified data will be provided via a Microsoft Excel file.

## Authors’ Contributions

Conceptualization: MA, EFF, HX

Data curation: MA, AN, HX

Formal analyses: AN

Funding acquisition: EFF, HX

Investigation: MA, HW

Methodology: EFF, AN, HX

Project administration: MA, EFF, HX

Resources: HX

Supervision: EFF, HX

Validation: EFF, HX, AN

Writing – original draft: MA

Writing – review & editing: MA, HW, EFF, AN, HX

## Abbreviations

IRB: Institutional Review Board
mYPAS: Modified Yale Preoperative Anxiety Scale
NCH: Nationwide Children’s Hospital
NRS: Numeric Rating Scale
PDL: Pulsed Dye Laser
PHI: Protected Health Information
RCT: Randomized Clinical Trial
REDCap: Research Electronic Data Capture
STAI-6: Short Form State Trait Anxiety Inventory
STAI-CH: State Trait Anxiety Inventory for Children
VR: Virtual Reality
VR-PAT: Virtual Reality Pain Alleviation Therapeutics

## Multimedia Appendix

SPIRIT Guidelines Checklist

---

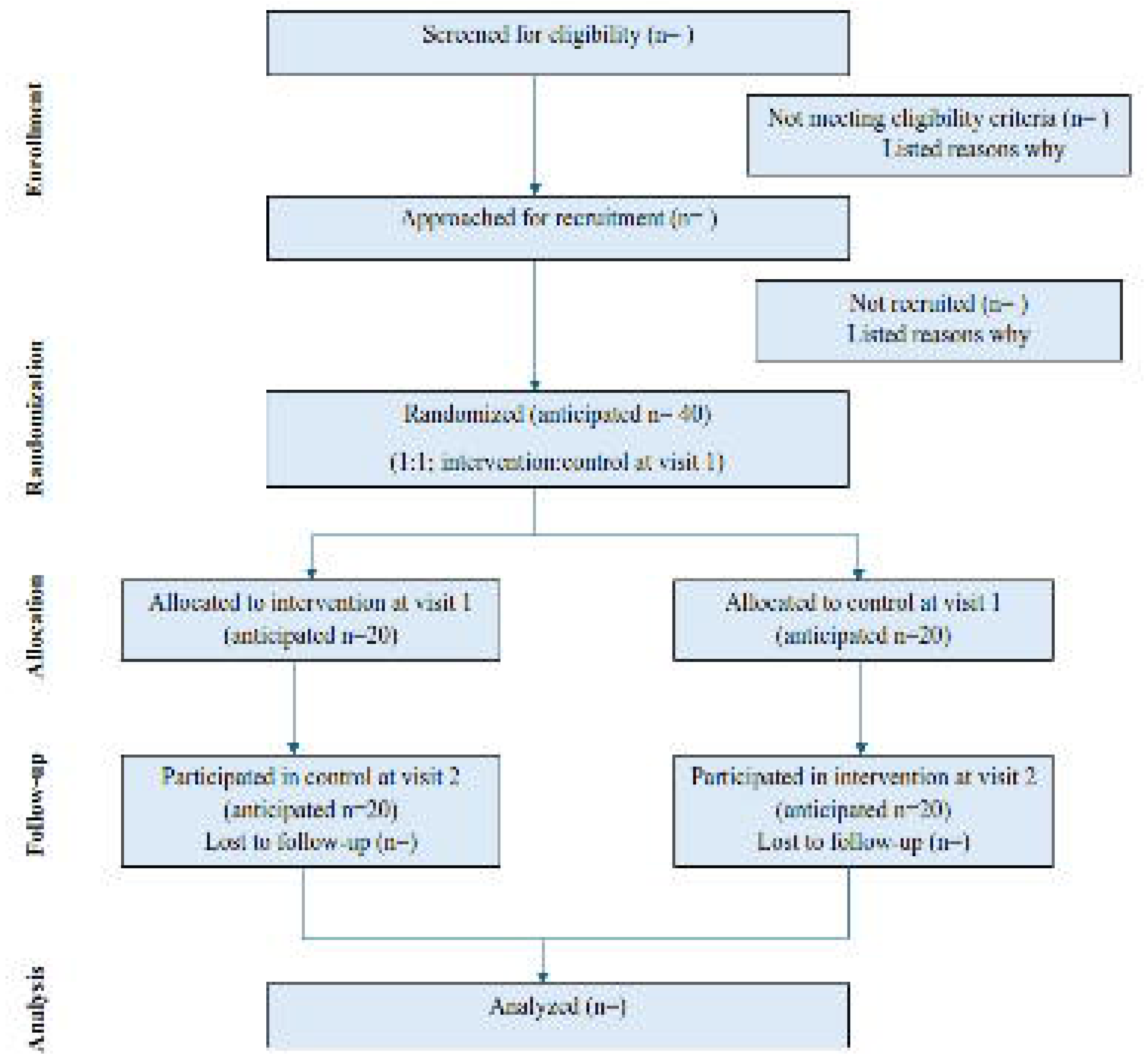

## References

1. Ducharme EE, Silverberg NB. Selected applications of technology in the pediatric dermatology office. Semin Cutan Med Surg. 2008 Mar;27(1):94–100. PMID: 18486031. doi: 10.1016/j.sder.2008.02.002.

2. Stevic M, Vlajkovic A, Trifunovic B, Rakic I, Ristic N, Budic I, et al. Topical anesthetics for pediatric laser treatment. J Cosmet Laser Ther. 2019;21(7-8):417–21. PMID: 31698962. doi: 10.1080/14764172.2019.1689273.

3. Cantatore JL, Kriegel DA. Laser surgery: an approach to the pediatric patient. J Am Acad Dermatol. 2004 Feb;50(2):165–84; quiz 85–8. PMID: 14726870. doi: 10.1016/j.jaad.2003.08.004.

4. Strauss RP, Resnick SD. Pulsed dye laser therapy for port-wine stains in children: psychosocial and ethical issues. J Pediatr. 1993 Apr;122(4):505–10. PMID: 8463892. doi: 10.1016/s0022-3476(05)83527-9.

5. Shahriari M, Makkar H, Finch J. Laser therapy in dermatology: Kids are not just little people. Clin Dermatol. 2015 Nov–Dec;33(6):681–6. PMID: 26686019. doi: 10.1016/j.clindermatol.2015.09.010.

6. Jenkins BN, Fortier MA, Kaplan SH, Mayes LC, Kain ZN. Development of a short version of the modified Yale Preoperative Anxiety Scale. Anesth Analg. 2014 Sep;119(3):643–50. PMID: 25010821. doi: 10.1213/ane.0000000000000350.

7. Kain ZN, Mayes LC, Caldwell-Andrews AA, Karas DE, McClain BC. Preoperative anxiety, postoperative pain, and behavioral recovery in young children undergoing surgery. Pediatrics. 2006 Aug;118(2):651–8. PMID: 16882820. doi: 10.1542/peds.2005-2920.

8. Nilsson S, Buchholz M, Thunberg G. Assessing Children’s Anxiety Using the Modified Short State-Trait Anxiety Inventory and Talking Mats: A Pilot Study. Nurs Res Pract. 2012;2012:932570. PMID: 22991660. doi: 10.1155/2012/932570.

9. Hildenbrand AK, Marsac ML, Daly BP, Chute D, Kassam-Adams N. Acute Pain and Posttraumatic Stress After Pediatric Injury. J Pediatr Psychol. 2016 Jan–Feb;41(1):98–107. PMID: 25825521. doi: 10.1093/jpepsy/jsv026.

10. Hoffman HG, Doctor JN, Patterson DR, Carrougher GJ, Furness TA, 3rd. Virtual reality as an adjunctive pain control during burn wound care in adolescent patients. Pain. 2000 Mar;85(1-2):305–9. PMID: 10692634. doi: 10.1016/s0304-3959(99)00275-4.

11. Nilsson S, Kokinsky E, Nilsson U, Sidenvall B, Enskär K. School-aged children’s experiences of postoperative music medicine on pain, distress, and anxiety. Paediatr Anaesth. 2009 Dec;19(12):1184–90. PMID: 19863741. doi: 10.1111/j.1460-9592.2009.03180.x.

12. Das DA, Grimmer KA, Sparnon AL, McRae SE, Thomas BH. The efficacy of playing a virtual reality game in modulating pain for children with acute burn injuries: a randomized controlled trial [ISRCTN87413556]. BMC Pediatr. 2005 Mar 3;5(1):1. PMID: 15745448. doi: 10.1186/1471-2431-5-1.

13. Loh TY, Cotton CH, Vasic JB, Goldberg GN. Current Practices in Pediatric Dermatology Laser Therapy: An International Survey. Lasers Surg Med. 2021 Sep;53(7):946–52. PMID: 32956533. doi: 10.1002/lsm.23327.

14. Bartels DD, McCann ME, Davidson AJ, Polaner DM, Whitlock EL, Bateman BT. Estimating pediatric general anesthesia exposure: Quantifying duration and risk. Paediatr Anaesth. 2018 Jun;28(6):520–7. PMID: 29722100. doi: 10.1111/pan.13391.

15. Nguyen CM, Yohn JJ, Huff C, Weston WL, Morelli JG. Facial port wine stains in childhood: prediction of the rate of improvement as a function of the age of the patient, size and location of the port wine stain and the number of treatments with the pulsed dye (585 nm) laser. Br J Dermatol. 1998 May;138(5):821–5. PMID: 9666828. doi: 10.1046/j.1365-2133.1998.02219.x.

16. Melzack R. From the gate to the neuromatrix. Pain. 1999 Aug;Suppl 6:S121–s6. PMID: 10491980. doi: 10.1016/s0304-3959(99)00145-1.

17. McCaul KD, Malott JM. Distraction and coping with pain. Psychol Bull. 1984 May;95(3):516–33. PMID: 6399756.

18. Wolitzky K, Fivush R, Zimand E, Hodges L, Rothbaum BO. Effectiveness of virtual reality distraction during a painful medical procedure in pediatric oncology patients. Psychology & Health. 2005 2005/12/01;20(6):817–24. doi: 10.1080/14768320500143339.

19. Hoffman HG, Richards TL, Bills AR, Van Oostrom T, Magula J, Seibel EJ, et al. Using FMRI to study the neural correlates of virtual reality analgesia. CNS Spectr. 2006 Jan;11(1):45–51. PMID: 16400255. doi: 10.1017/s1092852900024202.

20. Windich-Biermeier A, Sjoberg I, Dale JC, Eshelman D, Guzzetta CE. Effects of distraction on pain, fear, and distress during venous port access and venipuncture in children and adolescents with cancer. J Pediatr Oncol Nurs. 2007 Jan–Feb;24(1):8–19. PMID: 17185397. doi: 10.1177/1043454206296018.

21. Hoffman HG, Garcia-Palacios A, Kapa V, Beecher J, Sharar SR. Immersive Virtual Reality for Reducing Experimental Ischemic Pain. International Journal of Human–Computer Interaction. 2003 2003/06/01;15(3):469–86. doi: 10.1207/S15327590IJHC1503_10.

22. Hoffman HG, Patterson DR, Seibel E, Soltani M, Jewett-Leahy L, Sharar SR. Virtual reality pain control during burn wound debridement in the hydrotank. Clin J Pain. 2008 May;24(4):299–304. PMID: 18427228. doi: 10.1097/AJP.0b013e318164d2cc.

23. Schmitt YS, Hoffman HG, Blough DK, Patterson DR, Jensen MP, Soltani M, et al. A randomized, controlled trial of immersive virtual reality analgesia, during physical therapy for pediatric burns. Burns. 2011 Feb;37(1):61–8. PMID: 20692769. doi: 10.1016/j.burns.2010.07.007.

24. Gold JI, Kim SH, Kant AJ, Joseph MH, Rizzo AS. Effectiveness of virtual reality for pediatric pain distraction during i.v. placement. Cyberpsychol Behav. 2006 Apr;9(2):207–12. PMID: 16640481. doi: 10.1089/cpb.2006.9.207.

25. Carrougher GJ, Hoffman HG, Nakamura D, Lezotte D, Soltani M, Leahy L, et al. The effect of virtual reality on pain and range of motion in adults with burn injuries. J Burn Care Res. 2009 Sep–Oct;30(5):785–91. PMID: 19692911. doi: 10.1097/BCR.0b013e3181b485d3.

26. Hoffman HG, Richards TL, Coda B, Bills AR, Blough D, Richards AL, et al. Modulation of thermal pain-related brain activity with virtual reality: evidence from fMRI. Neuroreport. 2004 Jun 7;15(8):1245–8. PMID: 15167542. doi: 10.1097/01.wnr.0000127826.73576.91.

27. Hoffman HG, Patterson DR, Carrougher GJ, Sharar SR. Effectiveness of virtual reality-based pain control with multiple treatments. Clin J Pain. 2001 Sep;17(3):229–35. PMID: 11587113. doi: 10.1097/00002508-200109000-00007.

28. Dascal J, Reid M, IsHak WW, Spiegel B, Recacho J, Rosen B, et al. Virtual Reality and Medical Inpatients: A Systematic Review of Randomized, Controlled Trials. Innov Clin Neurosci. 2017 Jan–Feb;14(1-2):14–21. PMID: 28386517.

29. Xiang H, Shen J, Wheeler KK, Patterson J, Lever K, Armstrong M, et al. Efficacy of Smartphone Active and Passive Virtual Reality Distraction vs Standard Care on Burn Pain Among Pediatric Patients: A Randomized Clinical Trial. JAMA Netw Open. 2021 Jun 1;4(6):e2112082. PMID: 34152420. doi: 10.1001/jamanetworkopen.2021.12082.

30. Jaquez SD, Haller CN, England ME, Bruinsma RL, Arbet G, Croce EA, et al. Virtual reality and noise canceling headphone distraction during pediatric dermatologic procedures. Pediatr Dermatol. 2023 Nov–Dec;40(6):1161–3. PMID: 37816939. doi: 10.1111/pde.15401.

31. Armstrong M, Lun J, Groner JI, Thakkar RK, Fabia R, Noffsinger D, et al. Mobile phone virtual reality game for pediatric home burn dressing pain management: a randomized feasibility clinical trial. Pilot Feasibility Stud. 2022 Aug 18;8(1):186. PMID: 35982492. doi: 10.1186/s40814-022-01150-9.

32. Barnes LLB, Harp D, Jung WS. Reliability Generalization of Scores on the Spielberger State-Trait Anxiety Inventory. Educational and Psychological Measurement. 2002;62(4):603–18. doi: 10.1177/0013164402062004005.

33. Proczkowska M, Ericsson E. Validity of the modified-Distraction-Short-Scale and Verbal-Numeric-Anxiety-Fear-Rating-Scale for children in a preoperative setting. Paediatr Anaesth. 2024 Feb;34(2):121–9. PMID: 37728169. doi: 10.1111/pan.14765.

34. Castarlenas E, Jensen MP, von Baeyer CL, Miró J. Psychometric Properties of the Numerical Rating Scale to Assess Self-Reported Pain Intensity in Children and Adolescents: A Systematic Review. Clin J Pain. 2017 Apr;33(4):376–83. PMID: 27518484. doi: 10.1097/ajp.0000000000000406.

35. Castarlenas E, Sánchez-Rodríguez E, Vega Rde L, Roset R, Miró J. Agreement between verbal and electronic versions of the numerical rating scale (NRS-11) when used to assess pain intensity in adolescents. Clin J Pain. 2015 Mar;31(3):229–34. PMID: 24699160. doi: 10.1097/ajp.0000000000000104.

36. Marteau TM, Bekker H. The development of a six-item short-form of the state scale of the Spielberger State-Trait Anxiety Inventory (STAI). Br J Clin Psychol. 1992 Sep;31(3):301–6. PMID: 1393159. doi: 10.1111/j.2044-8260.1992.tb00997.x.

37. Kain ZN, Mayes LC, Cicchetti DV, Caramico LA, Spieker M, Nygren MM, et al. Measurement tool for preoperative anxiety in young children: The yale preoperative anxiety scale. Child Neuropsychology. 1995 1995/12/01;1(3):203–10. doi: 10.1080/09297049508400225.

38. Olsen MF, Bjerre E, Hansen MD, Hilden J, Landler NE, Tendal B, et al. Pain relief that matters to patients: systematic review of empirical studies assessing the minimum clinically important difference in acute pain. BMC Med. 2017 Feb 20;15(1):35. PMID: 28215182. doi: 10.1186/s12916-016-0775-3.

39. Tsze DS, Hirschfeld G, von Baeyer CL, Suarez LE, Dayan PS. Changes in Pain Score Associated With Clinically Meaningful Outcomes in Children With Acute Pain. Acad Emerg Med. 2019 Sep;26(9):1002–13. PMID: 30636350. doi: 10.1111/acem.13683.

40. Dai F, Silverman DG, Chelly JE, Li J, Belfer I, Qin L. Integration of pain score and morphine consumption in analgesic clinical studies. J Pain. 2013 Aug;14(8):767–77.e8. PMID: 23743256. doi: 10.1016/j.jpain.2013.04.004.

41. Sort R, Brorson S, Gögenur I, Hald LL, Nielsen JK, Salling N, et al. Peripheral nerve block anaesthesia and postoperative pain in acute ankle fracture surgery: the AnAnkle randomised trial. Br J Anaesth. 2021 Apr;126(4):881–8. PMID: 33546844. doi: 10.1016/j.bja.2020.12.037.

42. Jung MJ, Libaw JS, Ma K, Whitlock EL, Feiner JR, Sinskey JL. Pediatric Distraction on Induction of Anesthesia With Virtual Reality and Perioperative Anxiolysis: A Randomized Controlled Trial. Anesth Analg. 2021 Mar 1;132(3):798–806. PMID: 32618627. doi: 10.1213/ane.0000000000005004.

43. Girin H, Armstrong M, Bjorklund KA, Murphy C, Samora JB, Chang J, et al. Implementation of Virtual Reality Pain Alleviation Therapeutic into Routine Pediatric Clinical Care: Experiences and Perspectives of Stakeholders. J Med Ext Real. 2024 Sep;1(1):179–85. PMID: 39473565. doi: 10.1089/jmxr.2024.0018.

44. Maddox T, Sackman J, Stoudt M, Chibbaro M, Judge E, Rothery R, et al. Virtual reality-delivered skills-based therapy for chronic lower back pain: from proof-of-concept to FDA authorization to clinical implementation in-home and beyond. Frontiers in Virtual Reality. 2025 2025–June–24;Volume 6 - 2025. doi: 10.3389/frvir.2025.1613188.

